# Clinical Application of CT-Guided Lung Nodule Localization Needles in Preoperative Localization of Small Pulmonary Nodules

**DOI:** 10.64898/2026.04.13.26350830

**Authors:** Rongxue Xu, Hengli Dou, Meng Zhang, Zhenming Liu

## Abstract

**Background:** To investigate the safety and efficacy of CT-guided lung nodule localization needles for the preoperative localization of small pulmonary nodules.

**Methods:** A retrospective study was conducted on 102 patients with a total of 113 small pulmonary nodules who underwent preoperative localization at Jinan Fourth People’s Hospital from January 2024 to December 2025. Nodule diameter and depth, localization time, the number of pleural punctures, the localization success rate, and postoperative complications (hook dislodgement, hemorrhage, and pneumothorax) were recorded. All patients underwent video-assisted thoracoscopic surgery (VATS) after localization.

**Results:** The mean nodule diameter was 0.97±0.36 cm, the mean depth was 1.26±0.48 cm, and the mean localization time was 9.8±3.65 minutes. The hook dislodgement rate was 0.98% (1/102), the intrapulmonary hemorrhage rate was 14.71% (15/102), and the pneumothorax rate was 16.67% (17/102). All pulmonary nodules were successfully resected by VATS at 73.82±13.83 minutes after localization, and no severe complications occurred.

**Conclusions:** The use of a CT-guided lung nodule localization needle for the preoperative localization of small pulmonary nodules decreases the time needed for intraoperative nodule detection and operation time. This strategy is a simple, safe, and accurate preoperative localization method that is worthy of increased clinical use.

## Introduction

Lung cancer remains a common malignant tumor worldwide and has become the leading cause of cancer-related death in most countries. As lung tumors are the malignant tumors with the highest incidence and mortality globally, early diagnosis and treatment of lung cancer are crucial for improving patient prognosis. Accurate localization of small pulmonary nodules is key for early surgical treatment, given the extremely high incidence and mortality of this disease worldwide^12^. Therefore, early diagnosis and treatment of lung cancer are particularly important. With improvements in public health awareness and the need for physical examination, as well as the widespread application of high-resolution spiral CT, an increasing number of small pulmonary nodules have been detected^3^. Some small pulmonary nodules exhibit malignant biological behaviors; without timely intervention, these nodules may progress to malignant tumors, thereby affecting patient prognosis^4^. Video-assisted thoracoscopic surgery (VATS) has become the preferred surgical treatment for small pulmonary nodules in clinical practice because of its advantages, such as minimal invasiveness, rapid recovery, and the integration of diagnosis and treatment^56^. However, nodules located 1–2 cm or deeper below the pleura cannot be directly visualized under thoracoscopy. Small pulmonary nodules have a diameter of and are mostly ground-glass opacities, making accurate localization through palpation difficult or leading to instrument sliding^7^. Thus, preoperative localization is not only important but also challenging in thoracoscopic lung nodule surgery, as well as critical to the success rate of the operation and a reduced conversion rate to thoracotomy^89^. CT-guided percutaneous lung puncture localization has become the most commonly used preoperative localization method among clinical surgeons because of its advantages; this strategy is rapid and simple with a short learning curve, high resection success rate, and good patient acceptance^10^. In the past, a hook wire was used as a conventional preoperative localization method for small pulmonary nodules. Lung nodule localization needles are novel modified localization needles based on a hook wire; they are easy to use, low cost, and are associated with a low incidence of complications. Therefore, this study systematically analyzed the application of lung nodule localization needles.

## 1. Materials and Methods

### 1.1 Study Population

A retrospective analysis was performed on 102 patients with small pulmonary nodules (a total of 113 nodules) who underwent preoperative CT-guided lung nodule localization needle placement followed by VATS at Jinan Fourth People’s Hospital from January 2024 to December 2025. The inclusion criteria were as follows: (1) patients with small pulmonary nodules detected by high-resolution spiral CT with a diameter of ≤2 cm; (2) patients with nodules suspected of malignant potential on the basis of imaging features (e.g., ground-glass opacity with solid components, irregular shape, spiculation, or pleural indentation); (3) patients who voluntarily underwent preoperative localization and VATS and signed an informed consent form; and (4) patient with complete clinical data and follow-up records. The exclusion criteria were as follows: (1) patients with severe cardiopulmonary insufficiency, coagulation disorders, or other contraindications to puncture and surgery; (2) patients with multiple pulmonary nodules involving both lungs that could not be completely resected by VATS; (3) patients with metastatic tumors or other concurrent malignant tumors; and (4) patients with poor compliance or who were lost to follow-up.

### 1.2 Instruments and Equipment

The localization needle used was an SS510-10-type lung nodule localization needle (Ningbo Shengjiekang Biotechnology Co., Ltd.), which consists of a spherical spring coil, a three-color positioning marker, and a radiopaque ring, with a needle diameter of 21 G and a length of 10 cm.

The CT scanner used was a Philips 256-slice Brilliance iCT (Philips Healthcare, Netherlands) with a scan layer thickness of 1 mm and a resolution of 512×512 pixels. Other equipment included a disposable sterile puncture kit, 2% lidocaine hydrochloride injection, sterile gauze, a hemostatic forceps, and a positioning guide grid.

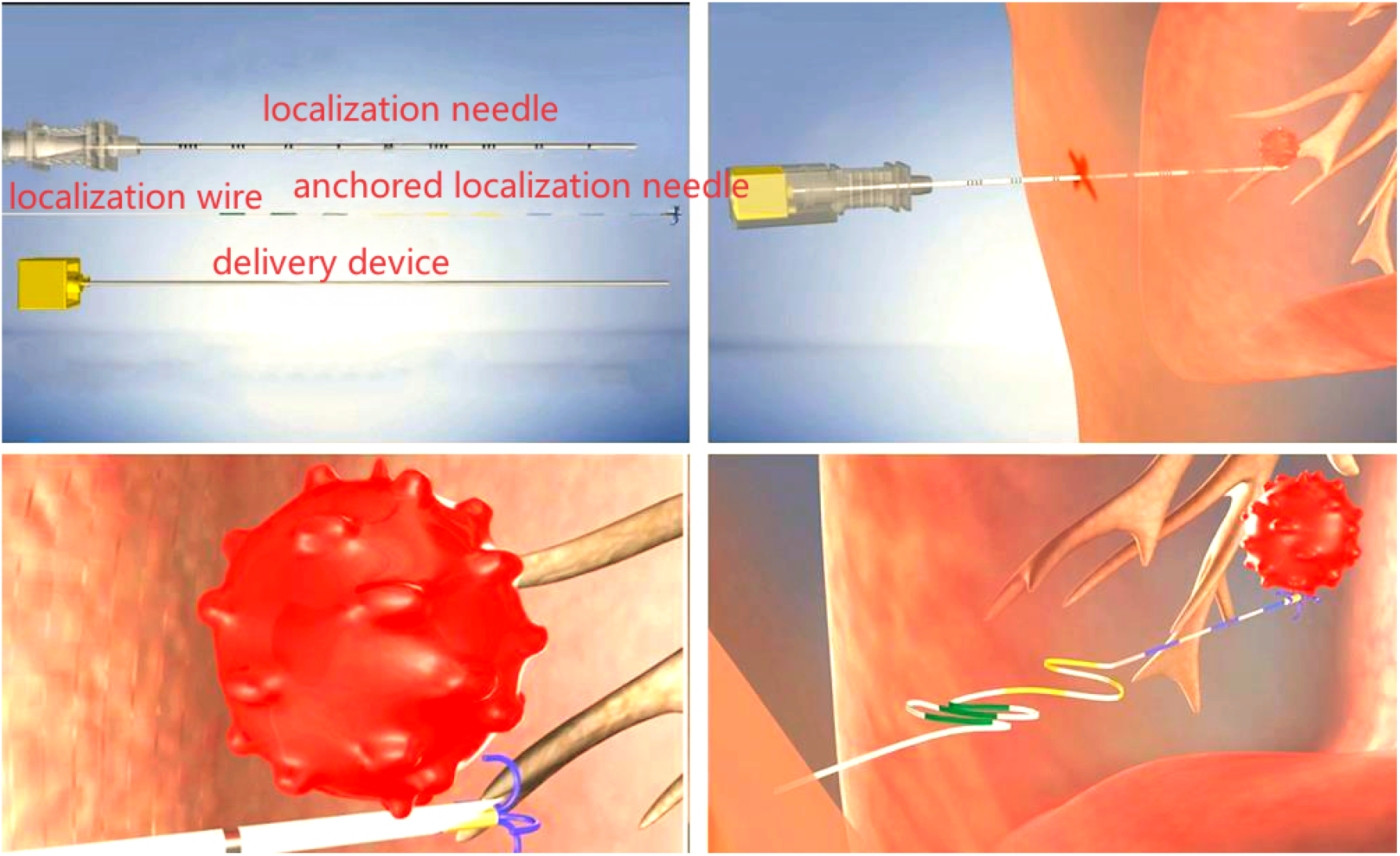

### 1.3 Localization and Surgical Methods

#### 1.3.1 Preoperative Preparation

All patients underwent routine preoperative examinations, including routine blood tests, coagulation function tests, electrocardiograms, and pulmonary function tests, to evaluate their surgical tolerance. Preoperative chest high-resolution CT was performed again to confirm the size, location, and depth of the nodules and their relationship with the surrounding tissues. Patients were instructed to practice breath-holding training to reduce respiratory movement during puncture.

#### 1.3.2 CT-Guided Localization Procedure

The patient was placed in a supine, prone, or lateral position according to the nodule location, and the puncture area was disinfected, after which sterile draping was applied. A positioning guide grid was placed on the chest wall corresponding to the nodule location, and a local CT scan was performed to determine the optimal puncture path (to avoid large blood vessels, bronchi, and ribs). Lidocaine hydrochloride (2%) was used for local infiltration anesthesia from the skin to the pleura. Under real-time CT guidance, the localization needle was advanced along the predetermined path, and the needle tip was adjusted to be 0.5–1.0 cm away from the nodule. After the correct position of the localization needle was confirmed by CT scan, the inner needle core was withdrawn, the spherical spring coil and three-color positioning marker were released, and the outer needle was slowly pulled out. A postlocalization CT scan was performed to check for complications such as pneumothorax or hemorrhage, and the patient was sent to the operating room for VATS within 2 hours.

#### 1.3.3 VATS Resection

Under general anesthesia with double-lumen endotracheal intubation, single-port VATS was performed through the 4th intercostal space of the anterior axillary line. The surgeon identified the three-color positioning marker and spherical spring coil under thoracoscopy and resected the nodule along with the surrounding 1–2 cm of normal lung tissue. The resected specimen was immediately sent for rapid freezing and pathological examination. If the pathological result indicated malignancy, further radical surgery (e.g., lobectomy or segmentectomy) was performed according to the tumor stage and patient condition.

**Figure 1.**
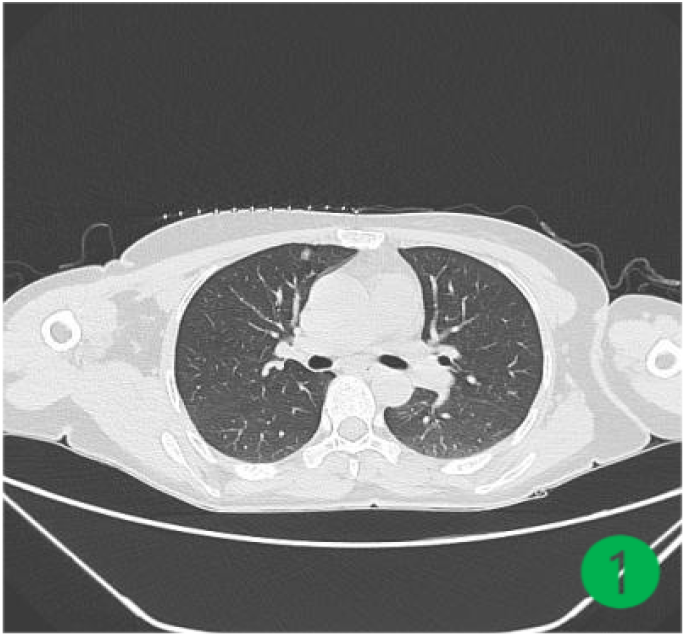
Positioning guide grid

**Figure 2.**
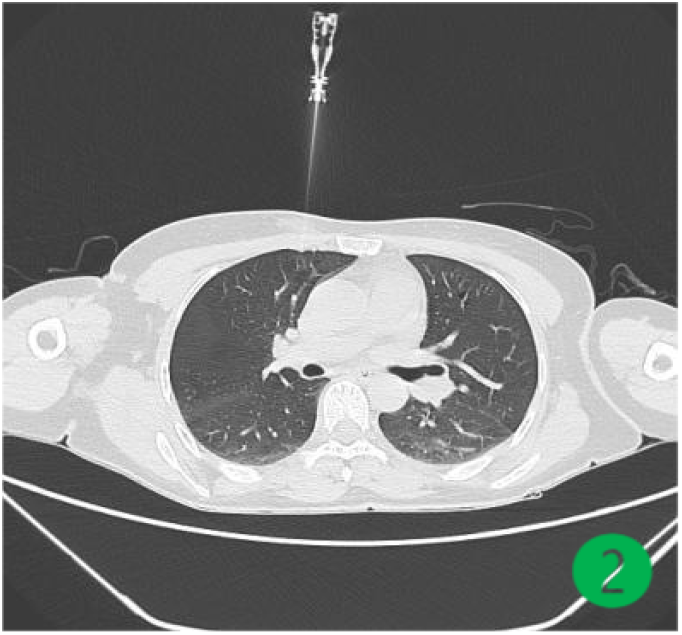
Localization needle

**Figure 3.**
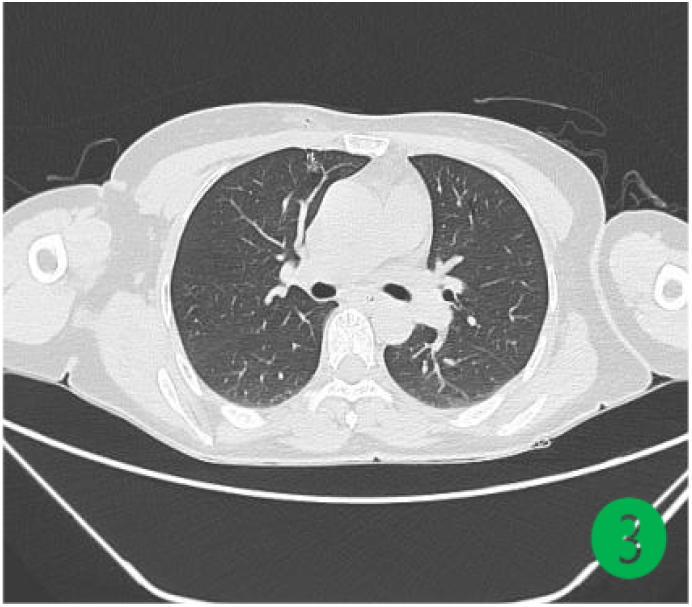
Anchored localization needle

**Figure 4.**
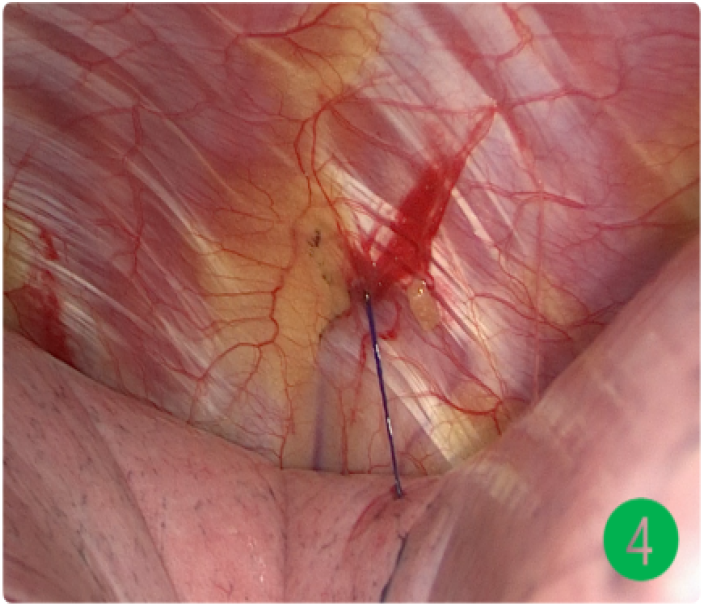
Localization wire

### 1.4 Observational Indicators

#### General indicators

General indicators included the nodule diameter (measured as the maximum diameter on CT images), nodule depth (the distance from the nodule center to the pleural surface), localization time (from the start of the CT scan to the completion of needle withdrawal), and number of pleural punctures.

#### Efficacy indicator

The localization success rate (defined as successful identification of the nodule during VATS and complete resection) was used as an efficiency indicator.

Safety indicators: Safety indicators included the incidence of complications, including hook dislodgement (loss of the positioning marker during surgery or CT examination), intrapulmonary hemorrhage (manifesting as pulmonary infiltration or hemoptysis on postlocalization CT), and pneumothorax (the need for chest tube drainage due to symptomatic pneumothorax or pneumothorax volume >30%).

#### Surgical indicator

The time from completion of localization to the start of VATS resection was used as a surgical indicator.

### 1.5 Statistical Analysis

Statistical analysis was performed using SPSS 26.0 software (IBM Corp., Armonk, NY, USA). Measurement data with a normal distribution are expressed as the mean ± standard deviation (x^−^±s), and comparisons between groups were conducted using independent samples t tests. Count data are presented as numbers (percentages) [n (%)], and comparisons between groups were performed using the chi-square test (χ^2^ test). A two-tailed P value of 5 was considered to indicate statistical significance.

## 2 Results

### 2.1 General Clinical Data of the Patients

**Table 1.**
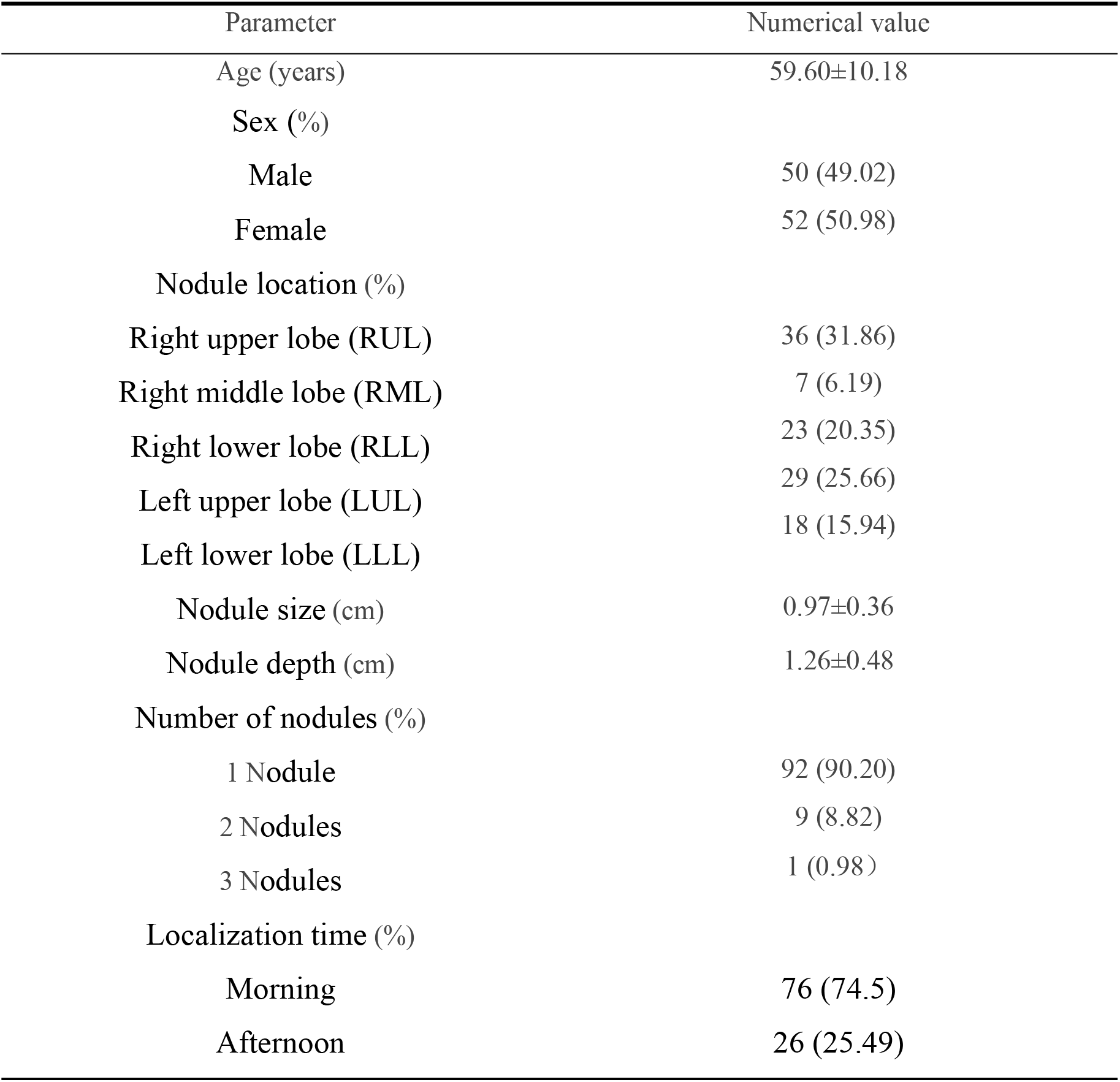
General Clinical Data of 102 Patients with Small Pulmonary Nodules.

| Parameter | Numerical value |
| --- | --- |
| Age (years) | 59.60±10.18 |
| Sex (%) |  |
| Male | 50 (49.02) |
| Female | 52 (50.98) |
| Nodule location (%) |  |
| Right upper lobe (RUL) | 36 (31.86) |
| Right middle lobe (RML) | 7 (6.19) |
| Right lower lobe (RLL) | 23 (20.35) |
| Left upper lobe (LUL) | 29 (25.66) |
| Left lower lobe (LLL) | 18 (15.94) |
| Nodule size (cm) | 0.97±0.36 |
| Nodule depth (cm) | 1.26±0.48 |
| Number of nodules (%) |  |
| 1 Nodule | 92 (90.20) |
| 2 Nodules | 9 (8.82) |
| 3 Nodules | 1 (0.98) |
| Localization time (%) |  |
| Morning | 76 (74.5) |
| Afternoon | 26 (25.49) |

The results revealed no statistically significant differences in patient age, sex, or localization time among the groups (all P > 0.05). In contrast, the nodule location and number of nodules were found to have statistically significant effects (both P values were 0.05). Specifically, the deeper the nodule location and the greater the number of nodules were, the higher the incidence of complications.

### 2.2 Localization Efficacy, Surgical Indicators and Complications

All the patients in this study underwent successful preoperative localization and subsequent VATS without any procedural failure. A total of 113 nodules were localized in 102 patients.

The key procedural and surgical indicators were as follows: the mean number of pleural punctures was 1.23±0.8; the average localization time was 9.8±3.65 minutes; the preoperative waiting time (from the completion of localization to the start of surgery) was 73.82±13.83 minutes; and the total operative time was 82.87±43 minutes.

In terms of postlocalization complications, the hook dislodgement rate was 0.98% (1/102), the incidence of intrapulmonary hemorrhage was 14.71% (15/102), and the pneumothorax rate was 16.67% (17/102). None of these complications required special treatment, and no severe adverse events occurred during the entire perioperative period.

**Table 2.** Complications of Small Pulmonary Nodule Localization.

| Complication | Incidence rate |
| --- | --- |
| Hook dislodgement (%) | 0.98% |
| Intrapulmonary hemorrhage (%) | 14.71% |
| Pneumothorax (%) | 16.67% |
| Overall complication (%) | 32.36% |

## 3 Discussion

Lung cancer is a malignant tumor with the highest incidence and mortality in China, and early and precise diagnosis and treatment are crucial for improving prognosis^11^. With increasing public health awareness and improvements in economic conditions, low-dose spiral computed tomography (LDCT) has been routinely used in health screenings. Consequently, the detection rate of small pulmonary nodules has increased exponentially. Among these nodules, some exhibit malignant biological behaviors and require further diagnosis and treatment. In traditional percutaneous lung biopsy and bronchoscopy, accurate determination of the benign or malignant nature of small pulmonary nodules is difficult. Therefore, VATS has become the optimal method to resect high-risk small pulmonary nodules. For micronodules, ground-glass nodules (GGNs), and nodules located 1–2 cm or deeper below the pleura, accurate identification of their specific positions during VATS through visual observation, finger palpation, or instrument clamping is challenging^12^. Clinicians often have to expand the resection range or convert to thoracotomy to improve the lesion resection rate. Thus, precise preoperative localization is particularly important^13,14^.

Precise preoperative localization can assist clinicians in formulating the optimal surgical plan, reducing trauma, maximizing the preservation of normal lung tissue, promoting early postoperative recovery, shortening hospital stay, and improving postoperative quality of life. This notion is consistent with the enhanced recovery after surgery (ERAS) management model^15^.

Hook wire localization is currently the most widely used percutaneous lung puncture localization method and was first developed by a team led by Professor Chen Haiquan from the Fudan University Shanghai Cancer Center^16^. A hook wire localization needle consists of two parts: the needle tip is a barbed needle (or metal hook) enclosed in a puncture trocar, which is used for fixation in lung tissue with an expanded length of approximately 1 cm, and the posterior part of the needle is a connecting wire approximately 30 cm long. Although the hook wire localization needle is the simplest and most practical type of localization device, it has a relatively high complication rate. The most common complication is pneumothorax, with an incidence rate of 35%, followed by intrapulmonary hemorrhage (15%). Additionally, hook dislodgement is a prominent issue, with relevant studies reporting an incidence rate of approximately 6%^10^. Severe complications such as air embolism^17^, tension pneumothorax, pleural reaction, and shock also occur occasionally. The pulmonary nodule localization needle used in this study is a novel modified needle based on the hook wire with the following characteristics: ① Localization anchor: a four-claw design ensures safety and firm fixation, and the anchor is not easy to dislodge (the hook dislodgement rate in this study was 0.98%); ② Spherical coil: no sharp structures are included, reducing damage from cutting and the risk of bleeding during localization (the pneumothorax rate and intrapulmonary hemorrhage rate in this study were 16.67% and 14.71%, respectively); ③ Three-color localization: the use of three-color localization makes it easier to identify the needle under VATS, allowing accurate measurement of insertion depth; ④ Radiopaque ring: the radiopaque ring allows clear visualization under CT, facilitating precise localization. All 102 patients in this study achieved successful localization without severe complications, and no special treatment was needed. All surgeries were performed without issue.

In addition to hook wire localization, other commonly used preoperative localization methods for pulmonary nodules in clinical practice include the following:① Percutaneous lung puncture with metal coil localization^18^: This technique is performed in essentially the same manner as hook wire localization. Commonly used metal coils include tornado microcoils, interlocking fibered coils, azur detachable coils, hybrid coils, and biodegradable coils that are currently under development, all of which show promising application prospects.② Percutaneous lung puncture with fiducial marker localization: This technique is advantageous because the path used to place fiducial markers does not affect the surgical approach and because localization and surgery can be performed at different times or even in different hospitals^19^. However, avoiding the placement of markers in veins and airways is crucial. A disadvantage of this technique is the requirement for intraoperative C-arm fluoroscopy to confirm the position of fiducial markers to ensure complete resection. ③ Percutaneous lung puncture with liquid marker injection: Methylene blue, the most commonly used medical dye, is inexpensive and easily accessible. However, its disadvantages include its rapid diffusion after injection and the requirement for surgery to be performed as soon as possible. Medical glue is a biomedical material with good biosecurity; after injection, it forms stable clumps in lung tissue that are not prone to peripheral diffusion and provide good tactile feedback during surgery, but medical glue may cause pain during injection. The combination of methylene blue and medical glue offers great advantages in the preoperative localization of pulmonary nodules and has achieved remarkable results^20^. ④ Electromagnetic navigation bronchoscopy (ENB)-assisted puncture localization: This technique minimizes the pain caused by CT-guided metal needle puncture localization and reduces specific complications such as pneumothorax and hemoptysis^21^, providing a safer option for the localization of multiple pulmonary nodules. However, this localization technique is overly dependent on computer processing, is complex to perform, and is associated with high costs^22^.

In summary, use of a CT-guided disposable pulmonary nodule localization needle is a safe, effective, simple, and economical technique for the preoperative localization of small pulmonary nodules with a low complication rate that is worthy of increased clinical application. However, this study has the following limitations: ① The sample size is relatively small, which may lead to potential bias, and ② the study is a single-center retrospective study lacking clinical controlled trials. Further large-sample, multicenter, prospective studies and comparative studies of various localization methods are needed to further verify the safety and effectiveness of pulmonary nodule localization needles. As pulmonary nodules are increasingly detected, the preoperative localization of small pulmonary nodules will undoubtedly become a top priority in minimally invasive thoracic surgery.

This study was approved by the Institutional Review Board (IRB) of Jinan Fourth People’s Hospital. Written informed consent was obtained from all patients or their legal representatives. All the authors declare that they have no conflicts of interest relevant to this article.

## Data Availability

The data used to support the findings of this study are included within the article.

